# Integrating miRNAs and Bacterial DNA for Early Detection of Lung Cancer

**DOI:** 10.1101/2023.03.23.23287641

**Authors:** Jun Shen, Huifen Zhou, Pushpa Dhilipkannah, Ashtosh Sachdeva, Edward Pickering, Van K. Holden, Janaki Deepak, Nevins W. Todd, Sanford A Stass, Feng Jiang

**Affiliations:** Department of Pathology, University of Maryland School of Medicine, 10 S. Pine St. Baltimore, MD 21201, USA; Department of Medicine, University of Maryland School of Medicine, Baltimore, MD, USA

**Keywords:** diagnosis, early stage, lung cancer, plasma, biomarkers

## Abstract

**Purpose:** The early detection is crucial for improved outcomes in lung cancer, which remains a leading cause of cancer-erelated deaths. There is an urgent need for precise molecular biomarkers to diagnose early-stage lung cancer. To address this, we assessed the potential of integrating diverse molecular biomarkers across both plasma and sputum to improve the accuracy of diagnosis, given the heterogeneous nature of lung cancer arising from multifactorial molecular aberrations.

**Methods:** We utilized droplet digital PCR to quantify miRNAs in plasma and bacterial DNA in sputum collected from 114 lung cancer patients and 121 cancer-free smokers. The participants were randomly divided into a development cohort and a validation cohort. Logistic regression models with constrained parameters were employed to optimize a signature with the highest sensitivity and specificity for early detection of lung cancer.

**Results:** The individual plasma miRNAs and sputum bacterial biomarkers had sensitivities of 62%-71% and specificities of 61%-79% for diagnosing lung cancer. A panel of plasma miRNA or sputum bacterial biomarkers produced sensitivities of 79%-85% and specificities of 74%-82%. An integrated signature comprising two miRNAs in plasma and three bacterial biomarkers in sputum was developed in the development cohort, and it exhibited a higher sensitivity (87%) and specificity (89%) in comparison to individual biomarkers. The signature’s diagnostic value was confirmed in the validation cohort, regardless of tumor stage, histological type, and demographic factors.

**Conclusion:** The integration of miRNA and bacterial biomarkers across both plasma and sputum samples offered an effective approach for the diagnosis of lung cancer.

## Introduction

Lung cancer is a deadly disease, with the highest mortality rate of all cancers(Aberle et al., 2011). Non-small cell lung cancer (NSCLC) is the most common type of lung cancer, accounting for approximately 85% of all lung cancer cases. NSCLC, mainly consists of two histological types, adenocarcinoma (AC) and squamous cell carcinoma (SCC). The survival rate for NSCLC varies from 10% to 56% depending on the stage at diagnosis. As the prognosis for patients with lung cancer is closely linked to the tumor stage, detecting lung cancer at an early or curable stage can significantly reduce mortality rates (Aberle et al., 2011). The early detection of lung cancer in a large randomized trial using low-dose CT (LDCT) has revealed a 20% relative reduction in mortality as compared to chest X-rays (Aberle et al., 2011). However, LDCT is associated with over-diagnosis, excessive cost, and radiation exposure (Patz et al., 2014). The development of molecular biomarkers that can precisely diagnose early-stage NSCLC is required.

During tumor development, cancer cells undergo apoptosis and necrosis, and release tumor-associated molecules that can circulate in bloodstream. The circulating tumors-derived molecules in plasma provide noninvasive cancer biomarkers. MicroRNAs (miRNAs) are small molecules and the aberrations contribute to carcinogenesis (Croce and Calin, 2005). Using whole-transcriptome NGS, we have systematically and comprehensively characterized miRNA profiles of lung tumors (Gao et al., 2015; Lin et al., 2020; Ma et al., 2014). We have identified three miRNAs (miRs-126-3p, 205-5p, and 210-3p) as plasma biomarkers for NSCLC(Lin et al., 2017).

Sputum is another type of noninvasively obtained specimen that is secreted from bronchi and bronchioles of the lower respiratory tract. Since molecular changes detected in sputum could reflect those in low respiratory tract, sputum can be substituted for the lower-airway fluids (e.g., bronchoalveolar lavage) and surgical tissues, which are more invasively collected, for detection of lung cancer. It has been recently suggested that microbiome play crucial roles in tumor formation via direct mutagenesis, promoting survival of tumor-initiating cells, or inducing pro-tumorigenic immune responses(Garrett, 2015) (Mao et al., 2018). Of the microbes, bacteria have been studied in malignancies, including lung cancer (Glyn and Purcell, 2022). We previously found that *Streptococcus* promoted lung tumorigenesis by triggering NF-kB pathways via binding PspC to PAFR(Li et al., 2023). We have recently shown that analysis of *Acidovorax, Capnocytophaga, Streptococus*, and *Veillonella* in sputum could improve the detection of NSCLC(Leng et al., 2021). Therefore, microbiota might provide biomarkers for the early detection and diagnosis of lung cancer.

Although the individual panels of plasma miRNA or sputum bacterial biomarkers show promise for lung cancer diagnosis, their sensitivities and specificities are not sufficient to be used in the laboratory settings. One of the major reasons might be that NSCLC is a heterogeneous disease driven by multifactorial molecular aberrations, only one type of molecular changes detected in one type of specimen may not achieve the diagnostic performance required in the clinics. In this study, we investigate if integrated analysis of the different types of molecular biomarkers across plasma and sputum can improve the early detection of lung cancer.

## Materials and Methods

### Patients and clinical specimens

Using a protocol approved by the University of Maryland Baltimore Institutional Review Board, we recruited lung cancer patients and cancer-free smokers according to the inclusion and/or exclusion criteria recommended by U.S. Preventive Services Task Force. Briefly, we enrolled smokers between the ages of 50-80 who had at least a 20 pack-year smoking history and were former smokers (quit within 15 years). Exclusion criteria included pregnancy, current pulmonary infection, surgery within 6 months, radiotherapy within 1 year, and life expectancy of < 1 year. The surgical-pathologic staging of NSCLC was used as the ground truth according to the TNM classification of the International Union Against Cancer (UICC) with the American Joint Committee on Cancer (AJCC) and the International Staging System for Lung Cancer.

We collected blood in BD Vacutainer spray-coated K2EDTA Tubes (BD, Franklin Lakes, NJ) and prepared plasma using the standard operating protocols developed by The NCI-Early Detection Research Network(Tuck et al., 2009). Briefly, the blood specimens were processed within one hour of collection by centrifugation at 1,300 X g for 10 minutes at 4°C. Sputum was collected from the participants before they received any treatment as described in our previous studies (Shen et al., 2014). The sputum samples were processed on ice in 4 volumes of 0.1% dithiothreitol (Sigma-Aldrich, St. Louis, Mo) followed by 4 volumes of phosphate-buffered saline (Sigma-Aldrich). We centrifuged the samples at 1500 x g for 15 minutes and removed the supernatant. The remaining cell pellets were collected and stored at -80°C until use.

### RNA isolation and droplet digital PCR (ddPCR) analysis of miRNAs

RNA was extracted from plasma by using Trizol LS reagent (Invitrogen Carlsbad, CA) and RNeasy Mini Kit (Qiagen, Hilden, Germany). The qualification and quantification of RNA were assessed by using Biospectrometer (Hutchinson Technology Inc, Hutchinson, MN) and Electrophoresis Bioanalyzer (Agilent Technologies, Foster City, CA). Reverse Transcriptase (RT) was carried out to generate cDNA by using a RT Kit (Applied Biosystems, Foster City, CA). ddPCR for analysis of expression level of miRNAs was performed as described in our published works by using a QX200™ Droplet Digital™ PCR System (Bio-Rad, Hercules, CA) (Shen et al., 2014; Shen et al., 2011). Briefly, PCR reaction mix containing cDNA was partitioned into aqueous droplets in oil via the QX100 Droplet Generator, and then transferred to a 96-well PCR plate. A two-step thermocycling protocol (95°C ×10min; 40 cycles of [94°C ×30s, 60°C ×60s], 98°C ×10 min) was undertaken in a T100™ thermal cycler (Bio-Rad). The PCR plate was then transferred to the QX100 Droplet Reader for automatic reading of samples in all wells. Copy number of each gene per µl PCR reaction was directly determined. We used QuantaSoft 1.7.4 analysis software (Bio-Rad) and Poisson statistics to compute droplet concentrations (copies/μL). Only miRNAs that had at least 10,000 droplets were considered to be robustly detectable by ddPCR in plasma and subsequently underwent further analysis (Ma et al., 2013). All assays were done in triplicates, and one no-template control and two interplate controls were carried along in each experiment.

### DNA isolation and ddPCR analysis of bacterial abundances

We used QIAGEN-DNeasy Blood & Tissue Kit (Qiagen) to isolate DNA according to manufacturer’s instructions. We determined the purity by taking the optical density (OD) of the sample at 280 nm for protein concentration and at 260 nm for DNA concentration. The ratio OD260 /OD280 was calculated and DNA sample within the range of 1.6-2 was considered as pure. We measured DNA copy numbers of bacterial genera by using the QX100 Droplet Digital PCR System and 2× ddPCR Supermix (Bio-Rad) with a protocol developed in our previous studies with modification(Leng et al., 2021). To generate the droplets, we inserted 20 µL of PCR reaction and 70 µL of Droplet Generation oil for Probes (Bio-Rad) in an eight-well cartridge using a QX100 droplet generator (Bio-Rad). We then transferred 40 µL of the generated droplet emulsion in a 96-well PCR plate (Eppendorf, Hamburg, Germany). Amplification reaction was conducted in a T100™ thermal cycler (Bio-Rad) with the following conditions: initial denaturation at 95°C for 5 min followed by 35 cycles of 15s at 95.0°C, 30s at 55.3°C, 5 min at 4°C, and finally, 5 min at 90°C for signal stabilization. After thermal cycling, we transferred plates to a droplet reader (Bio-Rad). We used the software provided with the ddPCR system for data acquisition to calculate the concentration of target DNA in copies/µL from the fraction of positive reactions using Poisson distribution analysis.

### Statistical analysis

To determine sample size, we set the area under the curve (AUC) of H0 (the null hypothesis) at 0.5. H1 represented the alternative hypothesis. To have a high reproducibility with adequate precision, we required ≥28 subjects per group. With this sample size, we would have 85% power to detect an AUC of 0.75 at the 2% significance level. Therefore, the sample size in the two cohorts could have enough statistical power. Pearson’s correlation analysis was applied to assess relationship of miRNA expressions or bacterial abundances with demographic and clinical characteristics of the participants. AUCs were used to determine accuracy, sensitivity, and specificity of each gene. We used the highest Youden’s J index (sum of sensitivity and specificity−1) to set up corresponding cut-off value. Logistic regression models with constrained parameters as in LASSO were used to eliminate the irrelevant factors, develop composite panels of biomarkers, and optimize a signature with the highest sensitivity and specificity. To compare the signature and our previously developed plasma and sputum biomarker panels, we compared their AUCs to determine the sensitivity, specificity, and accuracy as previously described (Leng et al., 2017).

## 3. Results

### The demographic and clinical variables of cases and controls

A total of 114 NSCLC patients and 121 cancer-free smokers were recruited. Among the cancer patients, 39 patients were female and 75 were male. Fifty-six had stage I NSCLC, 33 with stage II, 15 with stage III, and 10 with stage IV. Sixty-five lung cancer patients were diagnosed with AC, while 49 with SCC. Of the cancer-free smokers, 39 patients were female and 75 were male. There were no significant differences of age, gender and smoking status between the NSCLC patients and cancer-free smokers. The cases and controls were randomly grouped into two cohorts: a development cohort and a validation cohort. The development cohort consisted of 58 lung cancer patients and 62 cancer-free smokers, while the validation cohort comprised 56 lung cancer patients and 59 cancer-free smokers. The demographic and clinical variables of the two cohorts are shown in Supplementary Tables 1-2.

### The diagnostic performance of plasma miRNA and sputum bacterial biomarkers in distinguishing between lung cancer patients and cancer-free controls

ddPCR analysis of miRNAs in plasma and bacteria in sputum of a development cohort generated at least 10,000 droplets in each well of the plates. Therefore, miRNA expression and bacterial DNA abundances could be ‘‘read’’ by ddPCR for their absolute quantification in the clinical samples. The plasma miRNAs displayed a different level in 58 NSCLC patients compared with 62 control individuals (all p≤0.001) (Fig.1A) (Table 1). As a result, the individual plasma miRNAs had an AUC of 0.70 -0.81, producing 68.97% to 77.59% sensitivities and 66.63% to 78.69% specificities for detection of NSCLC (Table 1). Furthermore, the plasma miRNA biomarkers were significantly associated with AC (All p ≤0.05) (Fig.1A). In addition, combined use of the three plasma miRNAs as a panel of biomarkers created 79.31% sensitivity, 85.48% specificity, and 82.50% accuracy for diagnosis of NSCLC (Table 2). Since the plasma miRNAs were associated with histological types, the panel of three plasma miRNA biomarkers had a higher diagnostic value for AC with 81.82% sensitivity, 85.48% specificity, and 84.21% accuracy compared with SCC (72.00% sensitivity, 83.87% specificity, and 80.46% accuracy, all p<0.05) (Table 2). However, the plasma miRNA biomarkers didn’t have relationship with stage of NSCLC, and age, race, and sex of the participants (All p ≥ 0.05), except the smoking history (p<0.05) (Supplementary Table 3). Altogether, the plasma miRNAs could have the potential as biomarkers for the early-stage NSCLC, particularly lung AC.

**Fig. 1.**
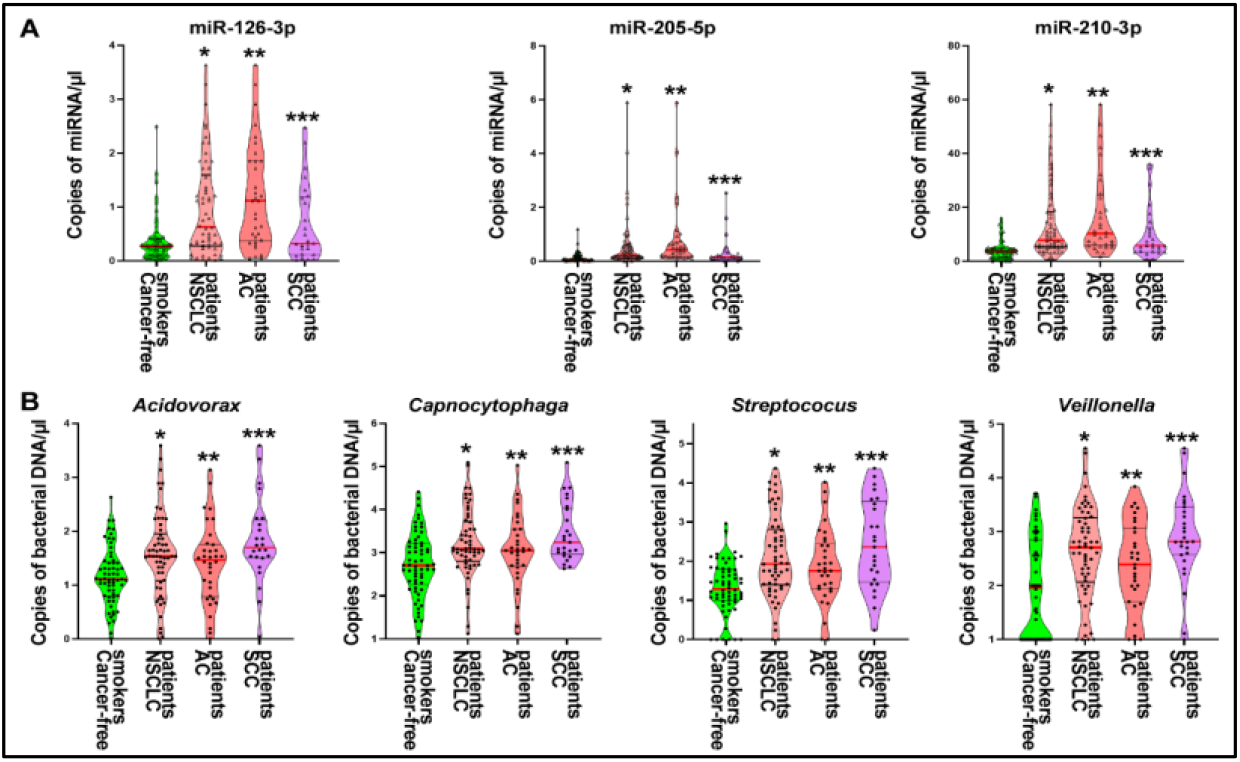
miRNAs in plasma and bacteria in sputum of 58 lung cancer patients and 62 controls. **A**. Plasma miRNAs displayed a higher level in NSCLC patients compared with control individuals (*, all p<0.05). The miRNAs show a higher level in AC patients compared with SCC patients (**, all p<0.05). The miRNAs show a higher level in SCC patients compared with cancer-free smokers (***, all p<0.05). **B**. Sputum bacteria genera have higher DNA abundances in NSCLC patients compared with control individuals (*, all p<0.05). Sputum bacteria genera display a higher level in AC patients compared with cancer-free smokers (**, all p<0.05). Sputum bacteria genera show a higher level in SCC patients compared with AC patients (***, all p<0.05).

**Table 1.** Plasma miRNAs and sputum bacteria genera in a development cohort of 58 lung cancer patients and 62 cancer-free smokers

| miRNAs or bacteria | Mean (SD) in cancer-free smokers | Mean (SD) NSCLC | P | AUC (95% CI) | Sensitivity % | Specificity % |
| --- | --- | --- | --- | --- | --- | --- |
| miR-126-3p | 0.269 (0.419) | 0.608 (1.393) | 0.0003 | 0.702 (0.606 to 0.798) | 68.97 | 66.63 |
| miR-205-5p | 0.0729 (0.183) | 0.2276 (1.013) | 0.0001 | 0.812 (0.735 to 0.889) | 69.26 | 82.26 |
| miR-210-3p | 3.700 (2.697) | 7.802 (13.689) | <0.001 | 0.806 (0.726 to 0.885) | 77.59 | 78.69 |
| *Acidovorax* | 1.106 (0.537) | 1.572 (0.786) | 0.0010 | 0.680 (0.581 to 0.779) | 62.07 | 79.03 |
| *Capnocytophaga* | 3.070 (0.957) | 3.667 (0.737) | 0.0009 | 0.656 (0.558 to 0.756) | 63.79 | 61.29 |
| *Streptococcus* | 1.312 (0.662) | 2.111 (1.065) | <0.0001 | 0.729 (0.639 to 0.820) | 70.86 | 54.29 |
| *Veillonella* | 1.010 (0.436) | 1.778 (0.737) | 0.0004 | 0.678 (0.579 to 0.776) | 68.97 | 58.06 |

Abbreviations: NSCLC, non-small cell lung cancer, pulmonary nodules; SD, standard deviation; AUC, the area under receiver operating characteristic.

**Table 2.**
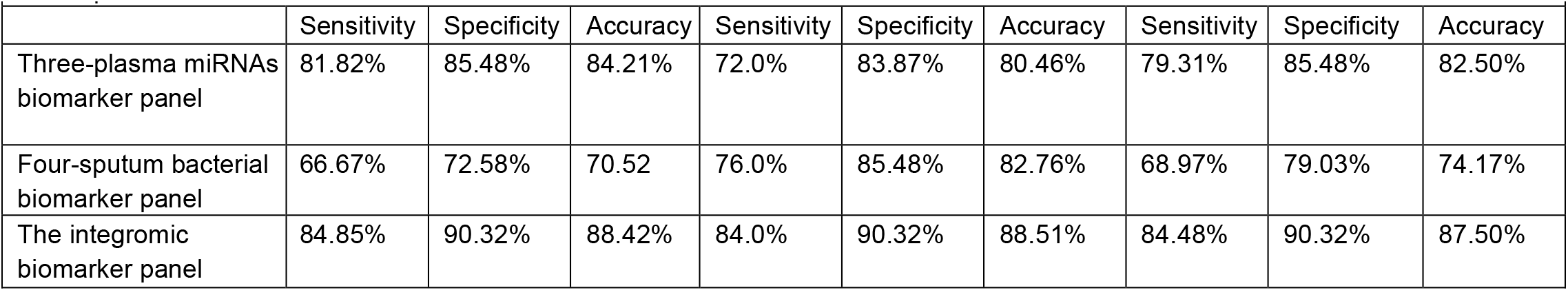
Diagnostic performance of individual plasma miRNA and sputum bacteria biomarker panel and integromic biomarker panel in development cohort

In the same discovery cohort, bacterial genera tested in sputum exhibited a higher level of DNA abundance in NSCLC patients compared with cancer-free smokers (All p≤0.001) (Fig.1B) (Table 1). The individual bacterial genera created 0.66-0.73 AUC with 62.07%-70.86% sensitivities and 61.29%-79.03% specificities for diagnosis of NSCLC (Table 1) (Supplemnetary Fig. 1). DNA abundance of the four bacterial genera in sputum was more related to SCC compared with AC (P<0.05) (Fig.1B). The use of the four bacteria genera in combination produced a hgiher AUC comapred with each bacteraim used alone (Supplemnetary Fig. 1). As a result, combined use of the four bacertia created 68.97% sensitivity, 79.03% specificity, and 74.17% accuracy for diagnosis of NSCLC, being significantly higher than each one used alone (Tables 1-2). The panel of sputum bacterial biomarkers had a higher diagnostic value for SCC with 76.00% sensitivity, 85.48% specificity, and 82.76% accuracy compared with AC (66.67% sensitivity, 72.58% specificity, and 70.52% accuracy, all p<0.05) (Table 2). The sputum bacterial biomarkers were not associated with stage of NSCLC, and age, sex, and ethnicity of the participants (All p ≥ 0.05), except the smoking history (p<0.05) (Supplementary Table 4). Therefore, the sputum bacterial biomarkers had the potential as biomarkers for the early-stage NSCLC, especially lung SCC.

### Combined use of plasma miRNA and sputum bacterial biomarkers for early detection of NSCLC

We further investigated if integrated analysis of the two classes of molecular changes in plasma and sputum might have a synergetic effect on the detection of lung cancer. We used least absolute shrinkage and selection operator (LASSO) to identify and optimize a panel of biomarkers for lung cancer. Two plasma miRNA (miRs-210-3p and -205-5p) and three sputum bacterial bacteria (*Acidovorax, Streptococus*, and *Veillonella*) biomarkers were selected as the best ones (all p<0.001) and integrated into a logistic model. Integrated use of the five biomarkers yielded a greater AUC (0.91) than did the three-plasma miRNA biomarker panel (0.87) or the four-sputum bacterial biomarker panel (0.79) (All p<0.05) (Fig.2). We used the highest Youden’s J index to set up corresponding cut-off value. The optimal cut-off for the integromic biomarker panel was U=1.27. Any subject with U≥1.27 was classified as a lung cancer case. As a result, the integromic biomarker panel produced significantly higher sensitivity (84.48%), specificity (90.32%), and accuracy (87.50%) compared with the individual panels of biomarkers (all p<0.05) (Table 2). The integromic biomarker panel could detect the positive cases that were positive by either plasma miRNA biomarkers or sputum bacterial biomarkers. Therefore, the integromic biomarker panel picked up the positive lung cancer cases identified by either the sputum biomarker panel or the plasma biomarker panel alone. Furthermore, integrated analysis of the diverse biomarkers across the different body fluids had no special association with stage and histology of lung cancer, and patients’ age, race, and gender (All P> 0.05).

**Fig. 2.**
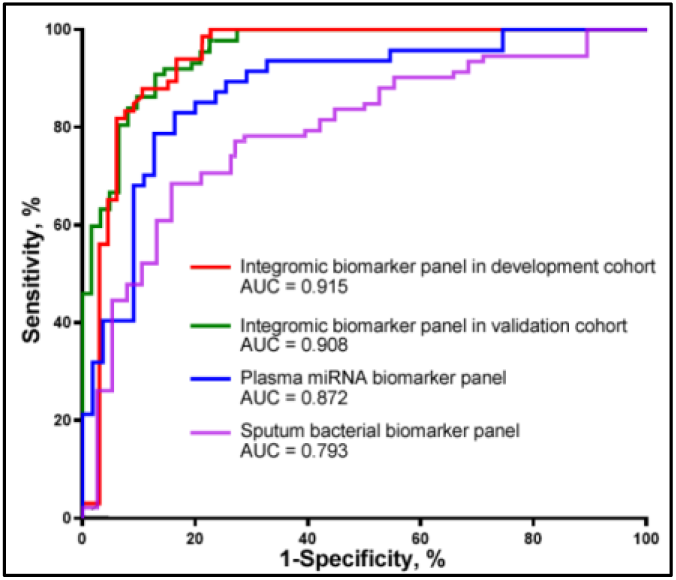
Plasma miRNA and sputum bacterial biomarkers for diagnosis of lung cancer. Blue line and purple line indicates diagnostic performance of three-plasma miRNA biomarker panel and four-sputum bacterial biomarker panel, respectively. Red line and green line shows the performance of the integromic biomarker panel in development cohort and validation set, respectively. There is no statistical difference of AUCs of the integromic biomarker panel in development cohort *vs*. validation cohort (p = 0.84).

### Validation of the integromic biomarker panel in a different cohort for the diagnosis of NSCLC

The three plasma miRNA and four sputum bacterial biomarkers included in the integromic biomarker panel were assessed in a validation cohort of 56 NSCLC patients and 59 cancer-free smokers. Combined analysis of the miRNAs and bacterial genera by using the logistic regression model created 0.90 AUC for lung cancer diagnosis. There was no significant difference between the development cohort and validation cohort regarding the AUCs (0.91 *vs*. 0.90, p=0.79) (Fig. 2). The biomarkers used in combination could differentiate the NSCLC patients from cancer-free smokers with 87.50% sensitivity, 89.83 % specificity, and 88.70% accuracy. In line with the findings in the development cohort, the integromic biomarker panel was independent of stage and subtype of NSCLC (all p>0.05). Moreover, there was no association of the molecular changes with the age, race, and gender (All p > 0.05), except smoking status of the individuals (p < 0.05).

## Discussion

Given that altered miRNAs and pathogenic bacteria contribute to tumorigenesis via the different mechanisms(Yang et al., 2022), we hypothesize that integrating the two types of molecular changes across the different body fluids may have a synergetic effect on the diagnosis of lung cancer. Our results show that the combined analysis of miRNAs in plasma and bacterial genera in sputum yielded a better performance than does a single category of the biomarkers, and thus validates the hypothesis. Furthermore, unlike plasma miRNA markers that are more specific to AC and sputum bacterial biomarkers that more specific to SCC, combined analysis of the plasma and sputum biomarkers is independent of histology of NSCLC, and hence substantiates the utility for predicting lung cancer. In addition, the integromic biomarker panel has a comparable diagnostic performance for lung tumor at the early versus late stages. Moreover, current screening tests lack effectiveness in detecting lung cancer in smoking populations, where the risk is higher. We results show that combining miRNAs in plasma and bacterial genera in sputum has superior diagnostic performance compared to using a single category of biomarkers. The discovery of the integromic biomarker panel would be important in utilizing it for the early detection of NSCLC in smokers.

Dysregulations of miRs-126-3p, 205-5p, and 210-3p have been found to associate with a variety of tumors, including NSCLC. For example, miR-126-3p can constrain cell proliferation of NSCLC *via* downregulating its target, EGFL7 (Shi et al., 2018). Our deep sequencing analyses have shown that miR-205-5p exhibits a higher expression level in lung tumor tissues compared with noncaseous lung tissues and could provide a potential biomarker for NSCLC (Li et al., 2017; Ma et al., 2014; Xing et al., 2010). Upregulation of miR-205-5p contribute to lung cancer cell proliferation and metastasis in *vitro* and in *vivo*, by regulating functions of TP53INP1, RB1, and P21(Zhao et al., 2022). Aberrant miR-210-3p expression in body fluids could diagnose several types of malignancies, including lung cancer (Papaconstantinou et al., 2013). Our current observations that the elevated expression levels of the three miRNAs in plasma of NSCLC patients support their important roles in lung carcinogenesis.

Among the bacterial genera analyzed, *Acidovorax* was found by Greathouse et al to have an elevated abundance in lung tumor tissues with TP53 mutation (Greathouse et al., 2018). Furthermore, there was a significant increase in lung tumor volume in mice inoculated with *Acidovorax. Acidovorax* could contribute to lung carcinogenesis in the presence of activated Kras and mutant p53, and thus act as a promoter in the development and progression of the disease (Greathouse et al., 2018). *Capnocytophaga* was proposed to be involved in lung carcinogenesis and lower respiratory tract infections (Yan et al., 2015). Furthermore, *Capnocytophaga* might induce long-term immune response/infection to the organ or cancer growth environment, which favors the growth of these bacteria in the airways (Gomes et al., 2019). We previously showed that *Streptococcus* attached to tumor cells by binding PspC to PAFR (Li et al., 2023). *Streptococcus* may have crucial function in carcinogenesis via triggering PI3K/AKT and NF-kB pathways. *Veillonella* was found to be overrepresented in lower airways of patients with NSCLC and related with ERK and PI3K signaling dysregulation (Tsay et al., 2018). Our current research suggests that *Veillonella* could serve as a potential biomarker for NSCLC. However, further studies are required to confirm the finding.

The study does have some limitations. 1), the sample size of the cohorts is small. We will perform a new study to prospectively validate the biomarkers for lung cancer early detection using a large population. 2), we only evaluate three miRNAs and four bacterial genera to develop this integromic biomarker panel. 87.50% sensitivity and 89.83% specificity of the integromic panel are not sufficient to be used in the clinics. To address the limitation, we are evaluating additional miRNAs and bacteria that are associated with NSCLC to develop more biomarkers to further improve the performance of the diagnostic approach. Furthermore, cell-free tumor DNA (ctDNA) or methylation profiles of ctDNA, have been suggested as potential biomarkers for lung cancer diagnostics and prognostics. We will compare the miRNA and bacterial DNA biomarkers with ctDNA biomarkers and investigate if their combination could provide a better diagnostic value for lung cancer.

In conclusion. Given the heterogeneous nature of NSCLC developed from multifactorial molecular aberrations, we have demonstrated that the integration of miRNA and bacterial biomarkers across plasma and sputum could provide an efficient approach for diagnosis of lung cancer. Nonetheless, a large multi-center clinical project to prospectively validate the full utility of the signature is required.

## Data Availability

All data produced in the present work are contained in the manuscript

## Author Contribution

JS, HZ, and FJ designed research approaches, conducted the experiments, and participated in data interpretation. PD, AS, ED, VH, JD, NT, SS consented patients and collected specimens. All authors read and approved the final manuscript.

## Data Availability

The data that support the findings of this study are available from the corresponding author upon a reasonable request.

## Ethics approval

The study was conducted according to the guidelines of the Declaration of Helsinki and approved by the Institutional Review Board of the University of Maryland Baltimore.

## Consent to participate

Informed consent was obtained from all subjects involved in the study.

## Consent to publish

The authors affirm that human research participants provided informed consent for publication of the resuts.

## Acknowledgements

This work was supported in part by the Geaton and JoAnn DeCesaris Family Foundation.

## Supplementary files

**Supplementary Table 1.** Characteristics of a development cohort of NSCLC patients and cancer-free smokers

|  | NSCLC cases (n = 58) | Controls (n = 62) | P-value |
| --- | --- | --- | --- |
| Age | 65.84 (SD 11.03) | 64.27 (SD 11.27) | 0.29 |
| Sex |  |  | 0.32 |
| Female | 20 | 23 |  |
| Male | 38 | 39 |  |
| Race |  |  | 0.34 |
| African Americans | 19 | 22 |  |
| White Americans | 39 | 40 |  |
| Smoking pack-years (median) | 33.7 | 30.5 | 0.13 |
| Stage |  |  |  |
| Stage I | 28 |  |  |
| Stage II | 17 |  |  |
| Stage III | 8 |  |  |
| Stage IV | 5 |  |  |
| Histological type |  |  |  |
| Adenocarcinoma | 33 |  |  |
| Squamous cell carcinoma | 25 |  |  |

Abbreviations: NSCLC, non-small cell lung cancer. SD, standard deviation.

**Supplementary Table 2.** Characteristics of a validation cohort of NSCLC patients and cancer-free smokers

|  | NSCLC cases (n = 56) | Controls (n = 59) | P-value |
| --- | --- | --- | --- |
| Age | 65.84 (SD 11.03) | 63.85 (SD 10.63) | 0.27 |
| Sex |  |  | 0.32 |
| Female | 19 | 23 |  |
| Male | 37 | 36 |  |
| Race |  |  | 0.36 |
| African Americans | 18 | 19 |  |
| White Americans | 38 | 40 |  |
| Smoking pack-years (median) | 34.8 | 31.9 | 0.19 |
| Stage |  |  |  |
| Stage I | 28 |  |  |
| Stage II | 16 |  |  |
| Stage III | 7 |  |  |
| Stage IV | 5 |  |  |
| Histological type |  |  |  |
| Adenocarcinoma | 32 |  |  |
| Squamous cell carcinoma | 24 |  |  |

Abbreviations: NSCLC, non-small cell lung cancer. SD, standard deviation.

**Supplementary Table 3.** The association of changes of miRNAs and bacterial genera with the age, gender, ethnic group, and tumor stage, and smoking status of the patients determined by Pearson’s correlation coefficient test. A p-value < 0.05 is statistically significant

|  | Age | Gender | Race | Smoking status | Stage |
| --- | --- | --- | --- | --- | --- |
| Genera | Correlation coefficients, P-value | Correlation coefficients, P-value | Correlation coefficients, P-value | Correlation coefficients, P-value | Correlation coefficients, P-value |
| *miR-126-3p* | -0.3486<br>0.3779 | -0.8437<br>0.2543 | -0.3087<br>0.7863 | 0.6520<br>**0.0344** | -0.7327<br>0.9953 |
| *miR-205-5p* | -0.4586<br>0.2868 | 0.2633<br>0.5413 | -0.3257<br>0.5853 | 0.4430<br>**0.0163** | -0.3237<br>0.2863 |
| *miR-210-3p* | -0.6216<br>0.7983 | -0.8663<br>0.5523 | 0.5653<br>0.2733 | 0.1050<br>**0.0289** | 0.2543<br>0.6903 |
| *Acidovorax* | -1.329<br>0.3893 | -0.1657<br>0.5623 | -0.4037<br>0.3643 | 0.7410<br>**0.03333** | -0.9127<br>0.3843 |
| *Capnocytophaga* | -0.4528<br>0.2883 | 0.2633<br>0.5413 | -0.3257<br>0.5853 | 0.4431<br>**0.0235** | -0.3237<br>0.2863 |
| *Streptococcus* | -0.5145<br>0.7563 | -0.4447<br>0.7763 | -0.6447<br>0.8873 | 0.6410<br>**0.0323** | -0.7237<br>0.7653 |
| *Veillonella* | -0.6632<br>0.1376 | -0.9657<br>0.2533 | -0.7537<br>0.2543 | 0.1217<br>**0.0355** | 0.9943<br>0.3643 |

**Supplementary Figure 1.**
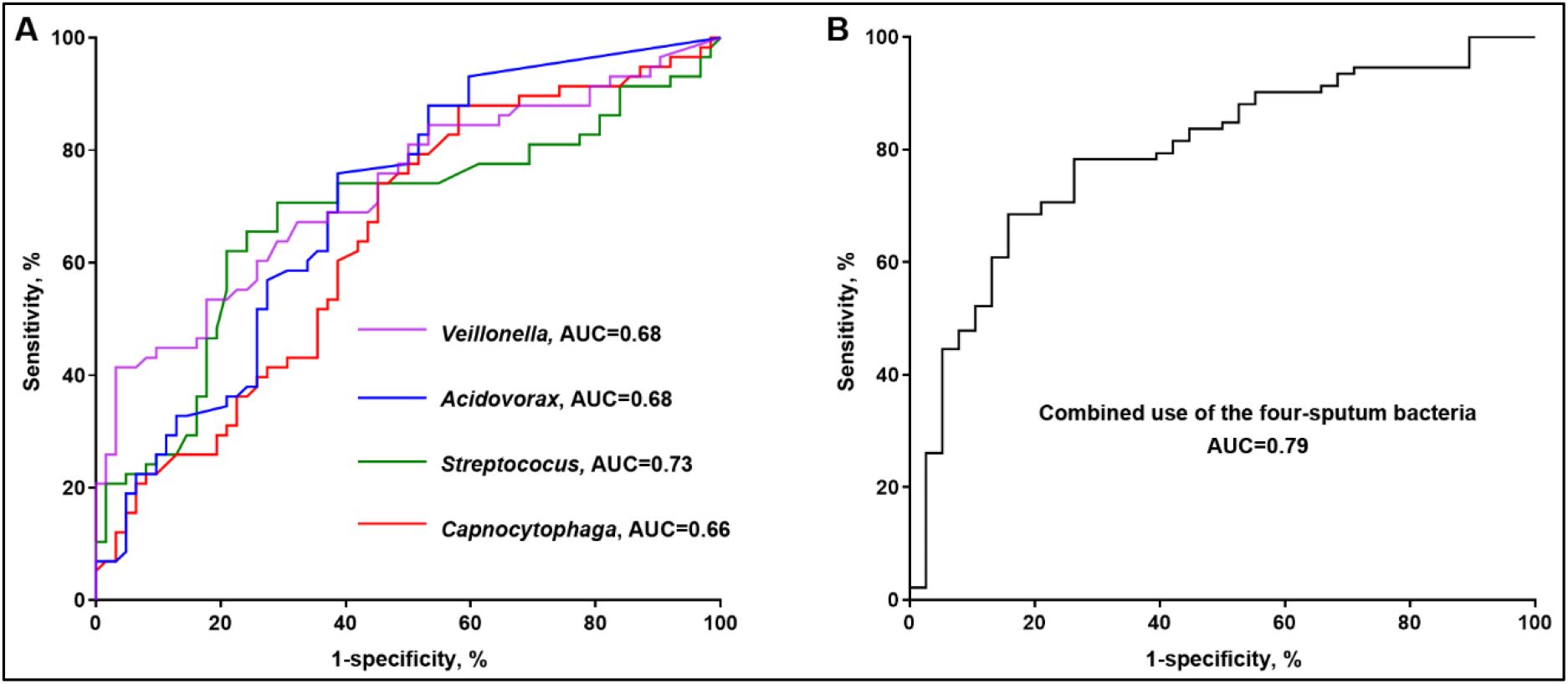
Combining four bacterial genera produced a higher AUC compared to each bacterium individually. ROC curve analysis was conducted on four bacterial genera in 58 lung cancer patients and 62 control individuals. The AUC values were calculated to assess the accuracy of each one in differentiating lung cancer patients from cancer-free individuals. **A**. Individually, the four bacterial genera produced AUC values of 0.66-0.73. **B**. When combined, the AUC value increased to 0.79, indicating higher accuracy in differentiating between the two groups.

